# Poolkeh Finds the Optimal Pooling Strategy for a Population-wide COVID-19 Testing (Israel, UK, and US as Test Cases)

**DOI:** 10.1101/2020.04.25.20079343

**Authors:** Yossi Eliaz, Mark Danovich, Gregory P. Gasic

## Abstract

The SARS-CoV-2 pandemic has changed the lifestyle of citizens of the world. In order for decision makers to manage the viral spread of COVID-19 both in the intra-national and the international frontiers, it is essential to operate based on data-driven assessments. It’s crucial to do so rapidly and frequently, since the nature of the viral spread grows exponentially and can burst worldwide again. A fast and accurate health status of individuals globally during a pandemic can save many lives and bring life back to a new normal. Herein, we present a data-driven tool to allow decision-makers to assess the spread of the virus among the world population. Our framework allows health agencies to maximize the throughput of COVID-19 tests among the world population by finding the best test pooling that fits the current SIR-D [1] status of the nation.

## 1 Introduction

As of today, there are more than 2.8 million confirmed COVID-19 cases in the world with a patient recovery rate of 27% (~ 800, 000). Sadly, due to COVID-19’s virulence more than 200, 000 (~7%) people have passed away while the rest are fighting for their lives. We have been witnessing how fast interventions by governments and decision makers save lives. Unfortunately, it isn’t trivial to test the whole population and it’s logistically hard. Quantitative RT-PCR (qPCR) tests are the most reliable option to test people for SARS-CoV-2. These tests have a high sensitivity and accuracy, and the machines used are available at major academic, biopharma, and commercial laboratories around the globe. Commonly, in order to detect a pathogen in individuals’ samples using qPCR, lab technicians need validated primers that target specific genes so the polymerase chain reaction will amplify only the genetic DNA of the virus to be detected. For RNA viruses qPCR relies on RNA purification, and calibration to determine not just the number of PCR DNA replication cycles that score as positive versus negative, but potentially viral load. Herein we report an easy to use, friendly, fast, accurate and mathematically solid platform to allow scientists and decision makers to plan the most efficient use of resources to test a large percentage of the population. As individuals begin to emerge from lockdown or mass isolation to resume more normal work and social activities, we must have a practical means to detect a resurgence of infections for contact tracing and/or return to isolation before symptomatic individuals begin to appear in our emergency rooms. Pooling of test samples for qPCR has the potential to save on limited reagents, resources, and time. In order to build the optimal test pooling, a measurement of the prevalence of the virus is in the population is required. Thus, we introduce a SIR-D pandemic model [1]. The idea of large population testing using bar-coded samples and next generation sequencing (NGS) has been proposed [2], as well as clinical proof that pooling [3, 4] using the most abundant COVID-19 test protocol is feasible. In order to find the best model for the viral spread in a specific country, we subject current epidemiological data to maximum-likelihood and the max-entropy principles. Then we project the prevalence of the viral infections to the near future and estimate the most stringently derived viral infection in a given population. Once we have the estimated prevalence in the population, we use a nested pooling strategy [5, 6] to find the optimal RNA pooling strategy for *n* individuals [7]. Alternatively, we can allow lab managers to find the best pooling or nested-pooling strategy of samples in order to screen the whole population expeditiously using any type of binary test. For example, if the COVID-19 prevalence in the population of 1 million people is 1%, using our approach for the whole population with 3-nested pooling steps, only 100,000 (10% of unpooled tests) actual tests need to be done.

## 2 Methods

### 2.1 The SIR-D differential equations and the equivalent time-varying Markov Chain

We incorporated a SIR-D model [1] to estimate the prevalence of the disease in the population. The SIR-D model considers four physically distinct measures: the number of individuals susceptible, infectious, recovered, and deceased people in a population. SIR-D can be written as four ordinary differential equations (ODEs) following Newton’s notation for time differentiation in Eq. (2). Here we allow the infection rate to decay with time as seen in Eq. (1). This should capture effects such as quarantine and other active interventions by the government. SIR-D is different than the conventional SIR model as it allows the infection rate *α*(*t*) to decay with time after *t*_0_ with decay rate *τ*^−1^ and defines an asymptotic infection rate *α*_0_*κ*.

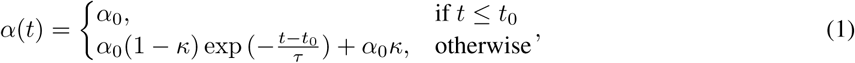

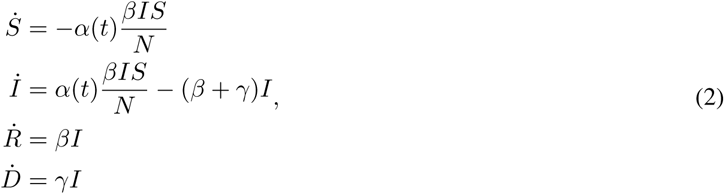

where *N* is the population size and *α*(*t*), *β*, and γ are the rates of infection, recovery and mortality, respectively. We note that as at any point in time we have *S* + *I* + *R* + *D* = *N*, it is therefore enough to know 3 of the 4 variables.

Moreover, the behavior of the viral infection spread can be modeled and simulated using a Markov chain with binomial distributions, as appears in Figure 1, and this is equivalent to the ODE model in Eq. (2). Using the maximum-likelihood principle and the Johns Hopkins dataset [8] we estimate the SIR-D parameters *τ*, *t*_0_, *к*, *α*_0_, *β*, and γ that maximize the log-likelihood at time *t*:

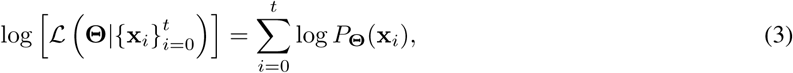

where x*_i_* represents the vector of the observed values: [*S*(*i*)*, I*(*i*)*, D*(*i*)] at time *i*, 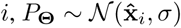 is the probability of these values to be x*_i_* at time *i*, assuming a normal distribution for the observed values with a mean 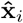 given by solving the SIR-D ODEs with the fitting parameters Θ = [*τ, t*_0_, к*, α*_0_*, β, γ*], and *σ* is the standard deviation of the distribution. 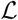 is the likelihood function which depends on Θ. By maximizing the likelihood in Eq. (3), we find the most probable SIR-D model parameters. Then we can simulate the progression of spread for a later time. For example, if we fit the data [8] for Israel, US, and the UK (Figure 2), until April 22, we can project the dynamics of COVID-19, and in this scenario we do it for 10-days.

**Figure 1:**
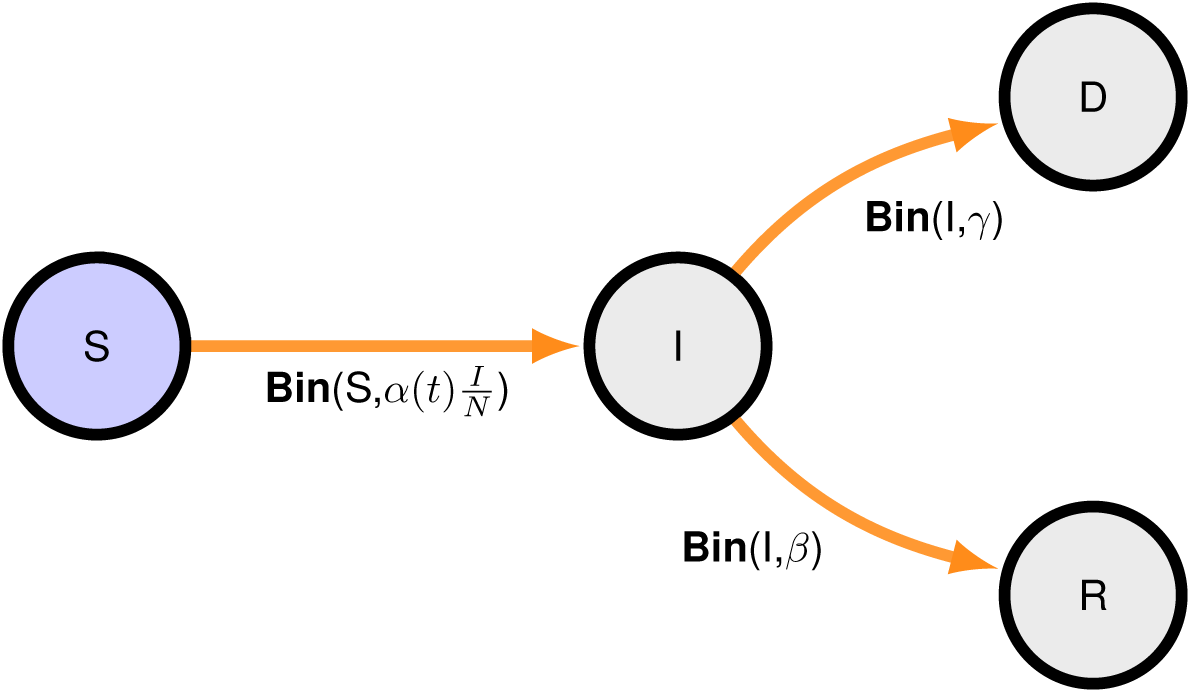
SIR-D as a time-varying Markov Chain. The notation **Bin**(*n, p*) corresponds the binomial distribution where *n* is the number of people in the prior state and *p* is the probability for an individual to move into the posterior state, e.g. 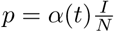 is the probability for an individual in the susceptible population of size *n* = *S* to get infected.

**Figure 2:**
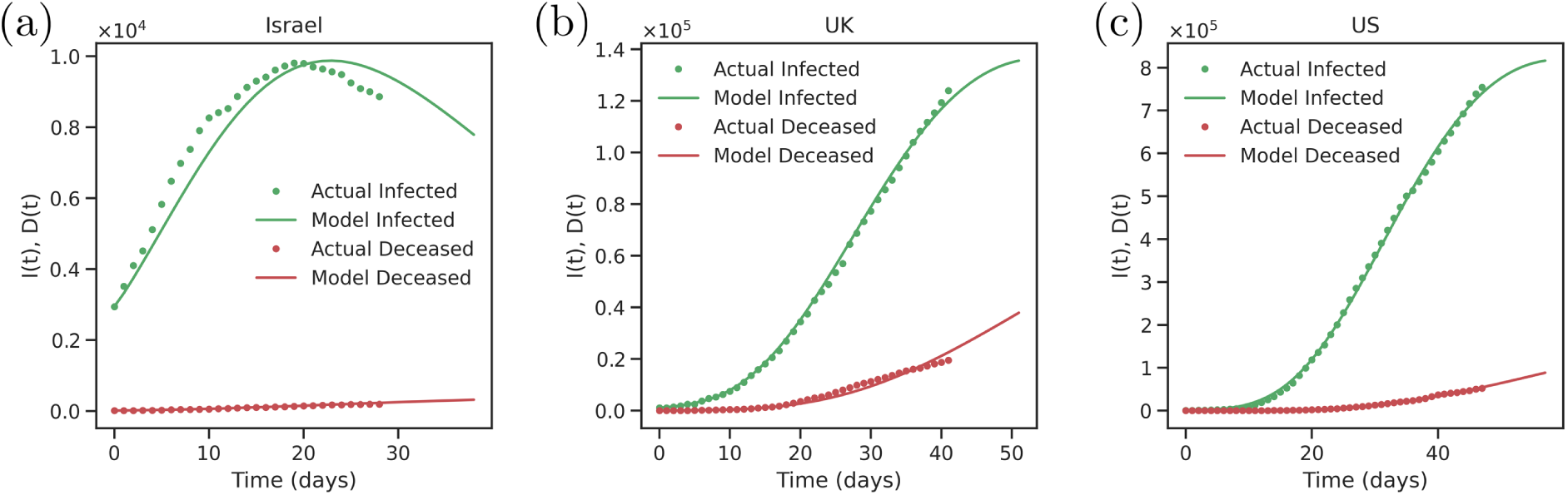
SIR-D prediction for the next 10-days from April 22, 2020 of *D*(*t*) and *I*(*t*) in (a) Israel, (b) US, and (c) UK.

### 2.2 Pooled Testing Strategy

Testing of multiple samples known as pooled testing [7] allows to significantly reduce the number of tests performed when screening a susceptible population, resulting in a quicker screening and using less resources as compared to screening each individual in the population. Once an estimate of the viral prevalence is available an optimal pooling strategy can be devised [6].

**Figure 3:**
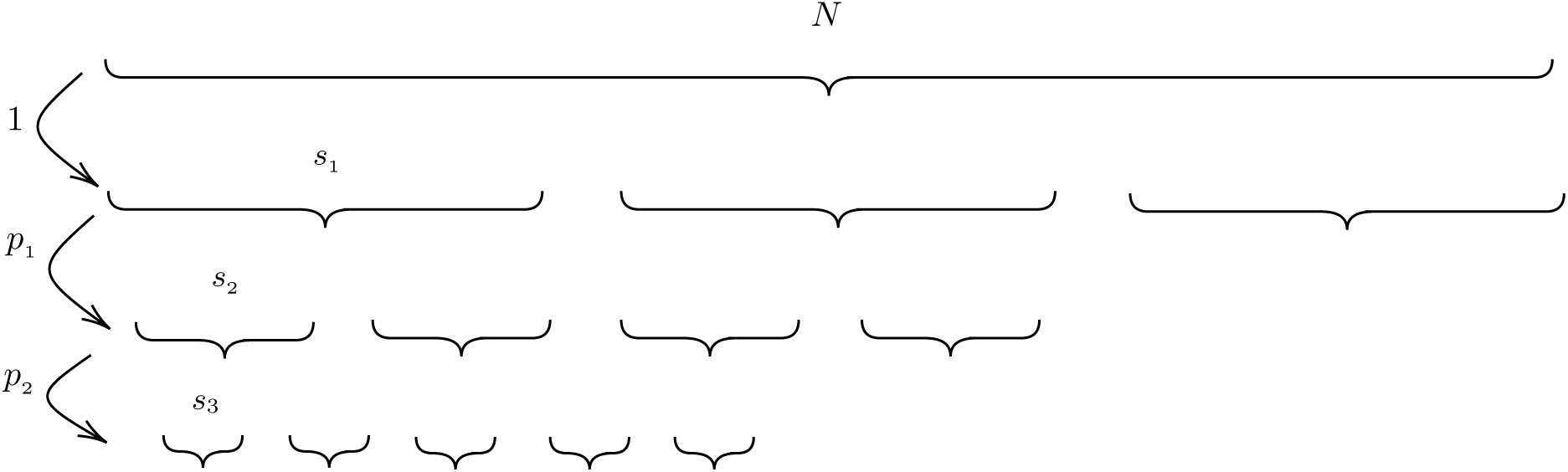
Nested pooling schematics. A population *N* is divided into pools containing *s*_1_, *s*_2_ and *s*_3_ samples per pool in each stage of the pooling. Each pooling stage reduces the population by a factor *p_i_* to a presumptive positive population used in the next stage of pooling. The final pooling consists of individual testing, *s*_3_ = 1.

### 2.3 Optimal Pooled Testing Strategy

In a population of size *N* with prevalence *r*_0_ of the virus the number of expected tests following a nested pooling strategy with *n* pools containing *s*_1_*, s*_2_*,.., s_n_* pools is given by

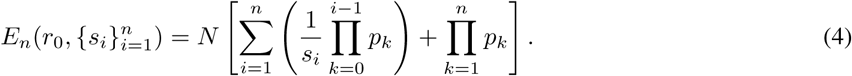

where 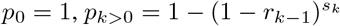 is the positive fraction in pool *k*, and 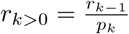 is the effective prevalence. The two most common examples in practice are the 1- and 2-pool testing. Using Eq. (4) with *n* = 1 and *n* = 2 we get,

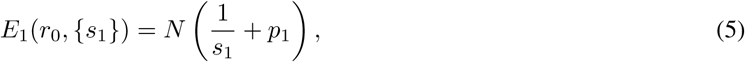

and,

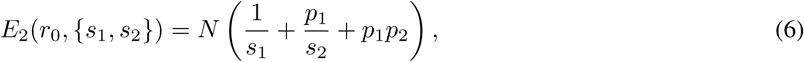

where 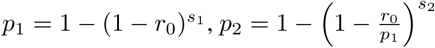.

We note that for a test with a non-zero false positive rate *ϕ*, the above expressions of *p_k_* are readily adjusted with the replacement below [5],

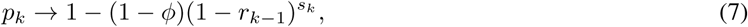

where (1-*ϕ*) is also known as the specificity of the test.

In Figure 4 we show the optimal pool sizes for 1-step and 2-step pool-testing and the expected number of total tests given as a percentage of the population *T* = *E_n_/N*. As Eq. (4) suggests, the pooling strategy is independent of the size of the population and the optimal strategy is derived only from the virus prevalence in the population.

**Figure 4:**
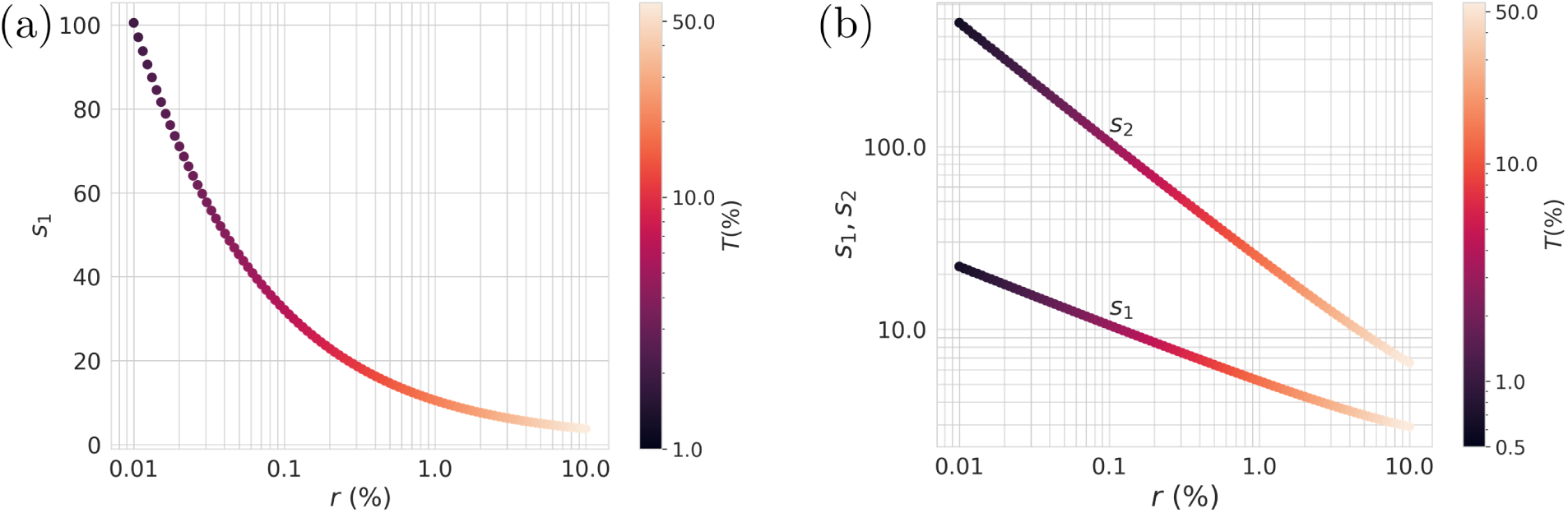
Simulation of the optimal sizes of the pools in (a) single-step pooling and (b) double-step pooling framework. The color bar of *T* represents the number of total performed tests (as percentage of the whole population) to screen the whole population. *r* is prevalence of the virus, and *s*_1_, *s*_2_ are the sizes of each pool in the first and the second steps, respectively.

**Figure 5:**
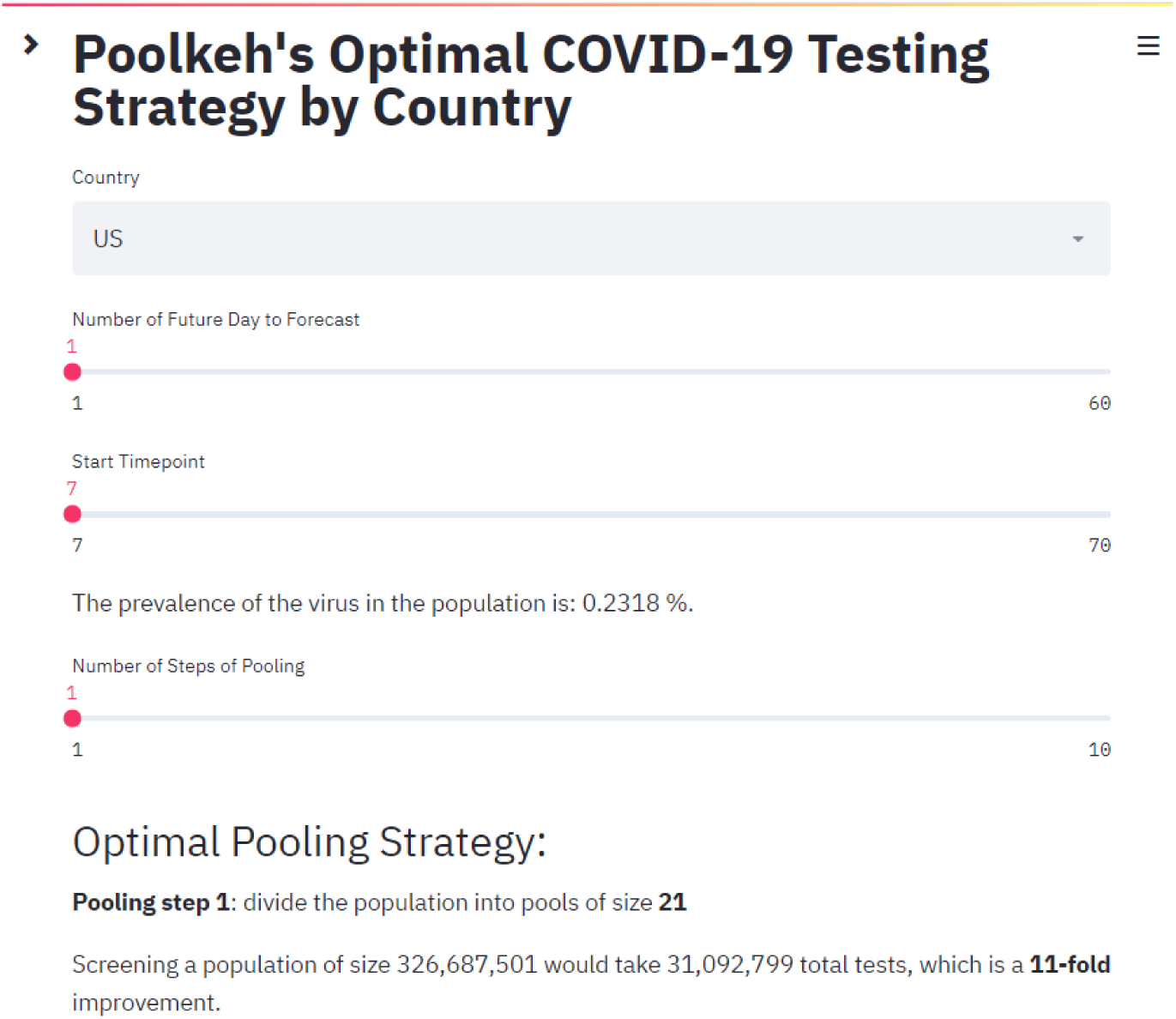
The User-Interface of Poolkeh-COVID-19 pooling optimizer.

## 3 Results

We are publishing Poolkeh as a free online tool at https://poolkeh.herokuapp.com/. Poolkeh has three pages and is accessible from the homepage (see Figure 6):

1. Poolkeh-COVID-19 allows the calculation of the optimal pooling strategy for a country.
2. Poolkeh also also allows the determination of the optimal strategy for pooling of test samples for any population of size *N* with prevalence of any pathogen *r* as shown in Figure 7.
3. Poolkeh-COVID-19 SIR-D predictor by country as depicted in Figure 5.

### 3.1 Poolkeh-COVID-19 for Israel, UK, and US

If we perform 2-step pooling of the entire state of Israel population (~ 9 million) we would need 295,951 tests with *s*_1_ = 92, and *s*_2_ = 10 which is 30-fold improvement over individual testing of the state of Israel.

If we perform 4-step pooling of the entire UK population (~ 69 million) we would need 2,300,000 tests with *s*_1_ = 159, *s*_2_ = 48, *s*_3_ = 14, and *s*_4_ = 4 which is 30-fold improvement over individual testing of the UK.

If we perform 5-step pooling of the entire US population (~ 330 million we would need 11,000,000 tests with *s*_1_ = 195, *s*_2_ = 73, *s*_3_ = 27, *s*_4_ = 10, and *s*_5_ = 3 which is more than 30-fold improvement over individual testing.

**Figure 6:**
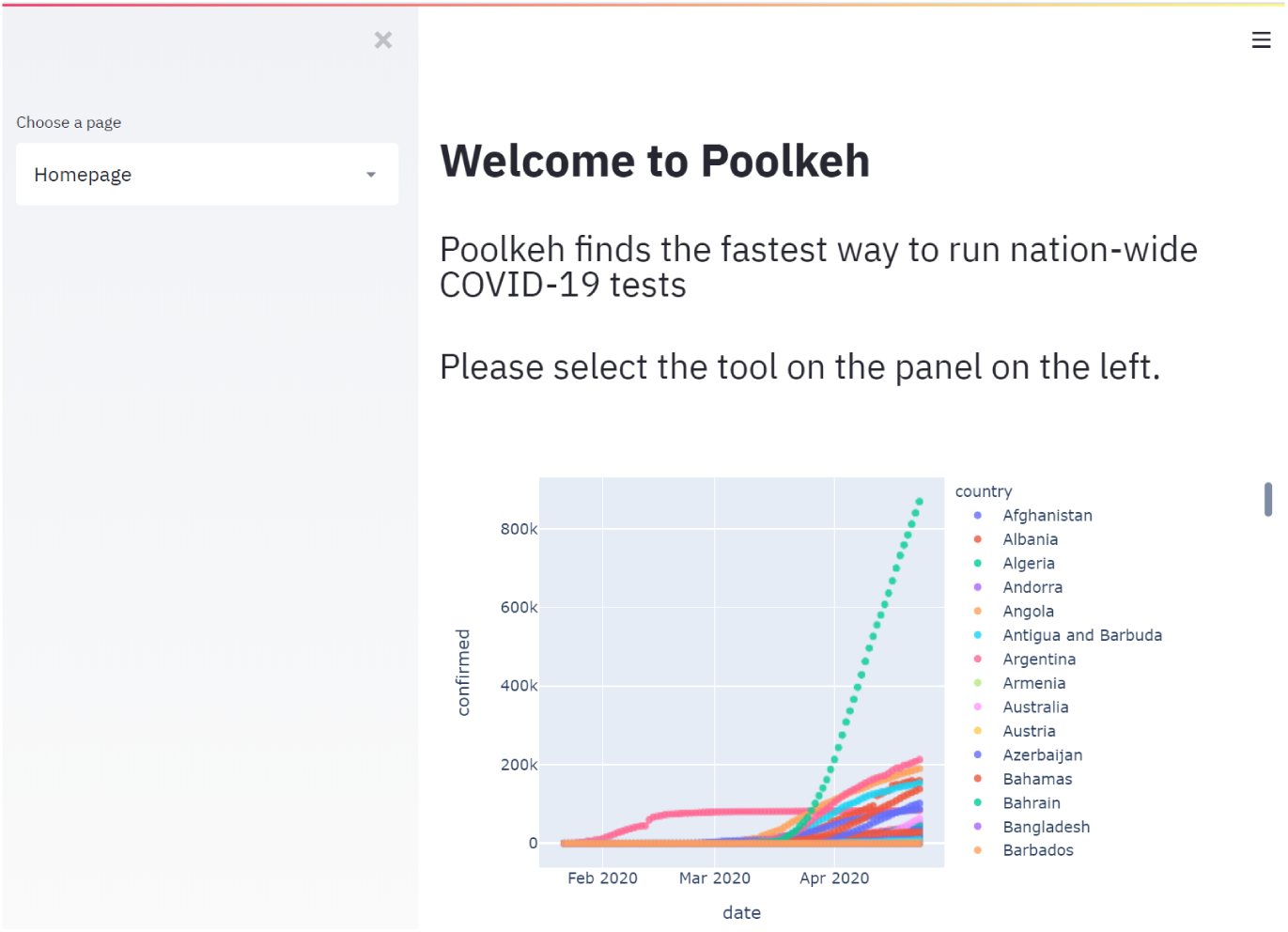
The Poolkeh homepage.

**Figure 7:**
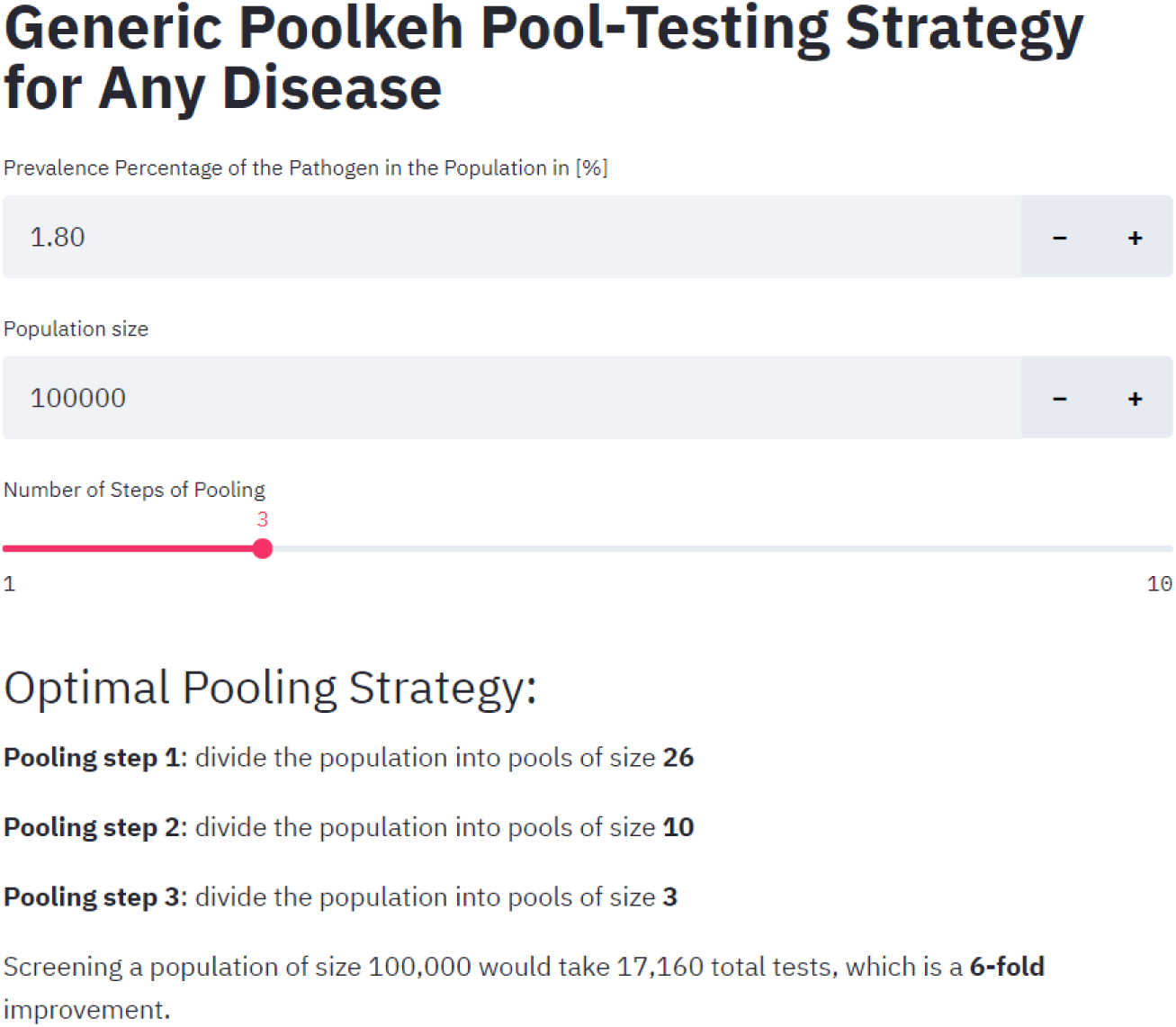
The Poolkeh Generic User-Interface.

## 4 Discussion

Poolkeh is an online tool, which allows users to develop an optimal scheme to pool tests for an entire population. The optimal strategy developed will depend on the prevalence of the virus in the test population, and the sensitivity of the test (qPCR or NGS). Nested pooling of RNA samples and qPCR will not only stretch the supply of resources (reagents) but also increase the estimation of viral prevalence at each testing time point as a larger percentage of the population is tested. Antibody testing has yielded a prevalence of infection between 2*−*30% of the population, but there may be pitfalls in the antibody tests and the population sampling [9]. A PCR approach with RNA pooling, however, can overcome some of the perils of relying on antibody test sensitivity and specificity. Moreover, viral immunity to SARS-CoV-2 is not the same as presence of IgG in an individual, and we still do not know whether the individual carries antibodies that neutralize and/or enhance viral infection for this virus. Although Real-time RT-PCR that uses individual amplification efficiencies for every PCR [10] is superior to the commonly used comparative *C_T_* (ΔΔ*C_T_*) and the standard curve method, a distributive pooling strategy of each individual sample into multiple amplification wells can provide a means to score each well as positive or negative. The amplified DNA copy number variation and efficiency of each amplification become less important. As shown recently [11], the prevalence of infection changes dramatically as actions by governments to isolate individuals that can spread the infection. Ideally, if time and resources allow it, a frequent population PCR-based tests [12] to obtain additional time points for viral infection prevalence would provide further insights into the potential number of tests necessary to update viral prevalence estimates. As the number of infections begins to drop and governments begin to impose less restrictive isolation measures to allow the normalization of economic and social activity, there needs to be an estimate of the number of tests required to determine that infection hot spots are not returning [13]. Incorporating daily data from an online nation-wide questionnaire [14] could also benefit the assessment of the prevalence. Regardless of how prevalence is computed, once available, Poolkeh finds the fastest nested test-pooling strategy without a lot of pipetting gymnastics. Moreover, Poolkeh isn’t restricted to RT-PCR, rather it can work with any clinical-proven testing techniques [2, 14]. While we were working on the last part of our manuscript, a preprint of a study with an interesting and thematically related approach [4] to testing for SARS-CoV-2 was posted and came to our attention.

## Data Availability

The data and tools that support the findings of this study are openly available in http://poolkeh.github.io/ and https://poolkeh.herokuapp.com/.

http://poolkeh.github.io/

## 5 Acknowledgements

We would like to thank three companies for their generosity: first to Netlify that gave us a free premium account to run static web applications on their servers, Heroku for their $700 credit to host our app, and finally, to Google which accepted our COVID-19 proposal and offered $600 in Google cloud computing credit. Poolkeh will run on Heroku’s credit in the next seven months, so labs around the world can optimize their pooling strategy for COVID-19 testing. We would like to express our sincere great hope for better and healthier times soon to come. Moreover, Y.E. thanks Professor Margaret S. Cheung, the Center for Theoretical Biological Physics (CTBP) at Rice University, and the University of Houston for their financial support via NSF PHY-1427654.

